# Accuracy of antigen and nucleic acid amplification testing on saliva and naopharyngeal samples for detection of SARS-CoV-2 in ambulatory care

**DOI:** 10.1101/2021.04.08.21255144

**Authors:** Solen Kernéis, Caroline Elie, Jacques Fourgeaud, Laure Choupeaux, Séverine Mercier Delarue, Marie-Laure Alby, Pierre Quentin, Juliette Pavie, Patricia Brazille, Marie Laure Néré, Marine Minier, Audrey Gabassi, Aurélien Gibaud, Sébastien Gauthier, Chrystel Leroy, Etienne Voirin-Mathieu, Claire Poyart, Michel Vidaud, Béatrice Parfait, Constance Delaugerre, Jean-Marc Tréluyer, Jérôme Le Goff

**Author notes:** **Corresponding author:** Solen Kernéis, Equipe de Prévention du Risque Infectieux, Hôpital Bichat, 46 rue Henri Huchard, 75018 Paris, France. Contributed equally (Solen Kernéis, Caroline Elie). Contributed equally (Constance Delaugerre, Jean-Marc Tréluyer, Jérôme Le Goff).

## Abstract

**Background:** Nasopharyngeal sampling for nucleic acid amplification testing (NAAT) is the current standard diagnostic test for of coronavirus disease 2019 (COVID-19). However, the NAAT technique is lengthy and nasopharyngeal sampling requires trained personnel. Saliva NAAT represents an interesting alternative but diagnostic performances vary widely between studies.

**Objective:** To assess the diagnostic accuracy of a nasopharyngeal point-of-care antigen (Ag) test and of saliva NAAT for detection of severe acute respiratory syndrome coronavirus-2 (SARS-CoV-2), as compared to nasopharyngeal NAAT.

**Design:** Prospective participant enrollment from 19 October through 18 December 2020.

**Setting:** Two community COVID-19 screening centers in Paris, France.

**Participants:** 1452 ambulatory children and adults referred for SARS-CoV-2 testing.

**Interventions:** NAAT on a saliva sample (performed with three different protocols for pre-processing, amplification and detection of SARS-CoV-2) and Ag testing on a nasopharyngeal sample.

**Measurements:** Performance of saliva NAAT and nasopharyngeal Ag testing.

**Results:** Overall, 129/1443 (9%) participants tested positive on nasopharyngeal NAAT (102/564 [18%] in symptomatic and 27/879 [3%] in asymptomatic participants). Sensitivity was of 94% (95% CI, 86% to 98%), 23% (CI, 14% to 35%), 94% (CI, 88% to 97%) and 96% (CI, 91% to 99%) for the nasopharyngeal Ag test and the three different protocols of saliva NAAT, respectively. Estimates of specificity were above 95% for all methods. Diagnostic accuracy was similar in symptomatic and asymptomatic individuals.

**Limitations:** Few children (n=122, 8%) were included.

**Conclusion:** In the ambulatory setting, diagnostic accuracy of nasopharyngeal Ag testing and of saliva NAAT seems similar to that of nasopharyngeal NAAT, subject to strict compliance with specific pre-processing and amplification protocols.

**Registration number:** NCT04578509

**Funding Sources:** French Ministry of Health and the Assistance Publique-Hôpitaux de Paris Foundation.

## Introduction

Prompt isolation of coronavirus disease-19 (COVID-19) cases is critical to mitigate the spread of the pandemic (1–3). This strategy implies rapid and reliable testing methods. Currently, the reference standard for diagnosis relies on detection of the severe acute respiratory syndrome coronavirus-2 (SARS-CoV-2) by nucleic acid amplification testing (NAAT) on a nasopharyngeal sample (NPS) (4,5). However, nasopharyngeal sampling must be performed by trained personnel, may be technically difficult in non-cooperating patients and requires specific sampling equipment. In addition, fear of discomfort and pain with nasopharyngeal swabbing discourage patients to attend testing. With NAAT testing, swabs are centralized in specialized virology laboratories and the technic itself requires around four hours to obtain results. Altogether, these constrains restrain access to massive testing, increase time-to-result and consequently delay isolation of infectious individuals (6).

Rapid point-of-care antigen (Ag) testing on NPS gives results in 15 to 30 minutes. Sensitivity of Ag tests was estimated at 50-90% and specificity at 90-100% as compared to nasopharyngeal NAAT (7,8). However, most evaluations were led on thawed SARS CoV-2-positive NPS diluted in viral transport media. Data are lacking on their point-of-care performances in real-life conditions, and in asymptomatic individuals.

Another strategy is to use alternative specimen types, such as saliva. Sample collection is simpler, painless, does not require specific skills of the personnel and opens the perspective of self-collection. Recent meta-analyses assessed diagnostic accuracies of saliva NAAT for the diagnosis of COVID-19 (9–12). All concluded that diagnostic accuracy was similar to that of nasopharyngeal swab NAAT, with pooled sensitivity estimates ranging from 83% to 88%. However, the literature review revealed great variations in specimen collection, processing protocols, populations included, and of sensitivity estimates of saliva NAAT. How this heterogeneity affects performances of the diagnostic strategies remains unknown. Indeed, most protocols were imprecise on specimen collection, criteria for interpretation of positive results. Selection procedures of participants were also marginally described. Additionally, data are lacking on asymptomatic individuals, and on evaluations led in real-life conditions, as part of routine screening in the community.

The objectives of this large prospective multicenter study were to compare diagnostic accuracy of two alternate diagnosis strategies (nasopharyngeal Ag test and saliva NAAT) to the current reference standard (nasopharyngeal NAAT) for detection of SARS-CoV-2 in community testing centers.

## Methods

### Setting

The study was conducted in two community screening centers located in Paris (France) within the COVISAN program (Assistance Publique-Hôpitaux de Paris, APHP). COVISAN is the regional declination of the national framework for testing and contact tracing in the French population. Briefly, all individuals with symptoms (i.e. temperature >37.8°C or chills, cough, rhinorrhea, muscle pain, loss of smell or taste, unusual persistent headaches or severe asthenia…) were invited to be tested for SARS-CoV-2 in one of the ambulatory COVISAN centers distributed across the region. Laboratory-confirmed cases were reached by phone by the COVISAN team within 24 hours to identify contacts as part of the contact tracing policy. Contacts were notified of their exposure within 24 hours of contact elicitation by phone. Symptomatic contacts were immediately referred for testing. Asymptomatic contacts were asked to self-quarantine and referred for testing 7 days after their last potential exposure. Apart from the contact tracing policy, testing was also available to all asymptomatic individuals wishing to be tested (i.e. before or after travel, participation to a gathering event).

### Study population and procedures

All adults and children, either symptomatic or asymptomatic, referred to the two participating COVISAN centers were eligible. Eligible persons received oral and written detailed information, adapted to their age. Participants were prospectively enrolled if they were not opposed to participate to the study. We collected data on sociodemographics, past medical history, presence of symptoms, consumption of alcohol, coffee, food, smoking and teeth brushing in the hours before testing. Participants were asked to evaluate nasopharyngeal and saliva sampling on a 0- to 10-point visual analog scale (VAS) for pain (0= no pain, 10= worst possible pain) and simplicity/convenience (0=not simple/convenient at all, 10=the most simple/convenient possible). For each participant, two NPS were collected by trained nurses. The first NPS was sent to the APHP high throughput platform for NAAT as part of routine care (reference method). The second NPS, collected in the second nostril, was used for rapid Ag testing immediately after sampling. The saliva sample was self-collected under supervision of the nurse, after nasopharyngeal swabbing. Saliva samples were centralized, frozen in several aliquots at -80°C within 24 hours and stored for analysis. As part of routine care, results of the nasopharyngeal NAAT were communicated to participants within 24 hours via a secured email. From 4 December 2020, following approval by health authorities of Ag testing in individuals with symptoms, Ag test results were disclosed to symptomatic participants immediately after testing (study protocol amendment number 3).

### Virology methods

#### Nasopharyngeal NAAT

NPS were centralized and processed according to the routine procedure (Appendix Table 1). Nucleic acid extraction was performed with MGIEasy Nucleic Acid Extraction Kit (MGI Tech Co, Shenzhen, China) on a MGISP-960 instrument (MGI Tech Co). SARS Cov-2 RNA amplification was done using TaqPath™ COVID 19 CE IVD RT PCR Kit (Thermo Fisher Scientific, Coutaboeuf, France). The technique provides results expressed as a cycle threshold (Ct) for each gene target (ORF1ab, N and S-genes).

#### Saliva NAAT

Saliva samples were tested with three NAAT procedures: “MGI-1”, “MGI-2” and “Roche” (Appendix Table 1). In the MGI-1 procedure, 150 µl of saliva was mixed with 500 µl of VSM02 buffer, incubated at 56°C for 30 min and then processed for extraction with the procedure used for nasopharyngeal NAAT. According to first examinations of data showing a poor sensitivity, sample testing with MGI-1 was discontinued and two alternative procedures set up (MGI-2 and Roche). All thawed saliva samples were extracted from the biobank and re-tested with the MGI-2 and Roche procedures. In the MGI-2 procedure, a 300 µl aliquot of saliva was mixed with 300 µl of NucliSENS^®^ lysis buffer (Biomerieux, Marcy l’Etoile, France) and extracted with the same procedure used for nasopharyngeal NAAT. In the Roche procedure, a 150 µL aliquot of saliva was mixed with 500 µL of Cobas omni Lysis Reagent and tested using the Roche cobas^®^ 6800 analyzer and Roche cobas^®^ SARS-CoV-2 assay (Roche Diagnostics France, Myelan, France) in the virology laboratory in Saint Louis hospital, Paris, France. The technique provides results expressed as a cycle threshold (Ct) for each gene target (ORF1ab and E-genes).

In addition, 93 other consecutive saliva samples were prospectively tested to compare fresh and frozen saliva using the MGI-2 method to detect SARS-CoV2 RNA (Appendix).

#### Rapid antigen testing

Nasopharyngeal Ag testing was performed with Standard Q COVID-19 Ag test (SD Biosensor^®^, Chuncheongbuk-do, Republic of Korea). Standard Q COVID-19 Ag test is a chromatographic immunoassay for the detection of SARS-CoV-2 nucleocapsid (N) antigen. The result was read after 15 to 30 minutes according to instructions of the manufacturer.

### Statistical Analysis

Sample size was calculated assuming that the sensitivity of the index tests was equal or superior to 60%. To allow sufficient precision (± 10%), 93 subjects with positive nasopharyngeal NAAT were needed in each of the two subgroups (symptomatic and asymptomatic participants). To account for samples excluded for technical reasons, a sample size of 110 subjects with positive nasopharyngeal NAAT was needed in each of these subgroups. With an average positivity rate of 8% in the participating centers, 2750 subjects needed to be included. On the planned meeting of the scientific committee of the study on 16 December 2020, considering that results were urgently needed in the context of rapid degradation of the epidemiological situation in France, the scientific committee recommended to perform the analysis on participants included up to 18 December 2020.

NAAT results were considered positive if at least one gene was detected : either N, S or ORF1ab (for MGI-1 and MGI-2), and either E or ORF1ab (for Roche). Analyses of tests results were carried out blind of the result of the others and of the participant’s clinical data. For each technique, Ct values reported are those for the ORF1a gene, and if not amplified, of the E-gene for Roche and of the N-gene for MG-1 and MG-2 (and of S-gene if the N-gene was not amplified).

Quantitative data were expressed as median [interquartile range], and qualitative data as counts (percentages). Diagnostic accuracy of the index tests was evaluated by calculating sensitivity and specificity. Confidence intervals were calculated by the exact binomial method. Subgroups analyses were performed according to: i) the presence of symptoms on day of testing, ii) the Ct value of the nasopharyngeal NAAT, expressed as low (at least one of the 3 targets with Ct ≤ 28, i.e. high viral shedding), or high (all 3 targets with Ct > 28, i.e. low viral shedding), and (iii) to the consumption of alcohol, coffee, food, and smoking or teeth brushing before sample collection.

Sensitivity analyses were performed considering 2 alternate criteria for positivity for the reference standard: i) ≥ 2 positive targets with nasopharyngeal NAAT, and ii) ≥ 1 positive target with either the nasopharyngeal NAAT, saliva MGI-2 or saliva Roche. The second sensitivity analysis was performed to address the fact that the nasopharyngeal NAAT represents an imperfect reference standard, as shown by others (10).

Quantitative variables were compared with Wilcoxon’s (paired test if appropriate) or Kruskal Wallis tests and qualitative variables Fisher’s exact tests (Mc Nemar test if appropriate). Correlations between Ct values of nasopharyngeal and saliva tests were assessed by calculating the Spearman’s correlation coefficient (*ρ*). Agreement between methods for Ct values was assessed by calculating intraclass correlation coefficients (ICC), and on Bland-Altman plots. All statistical tests were 2-sided with a significance level of 5%. The statistical analysis was performed using R software (http://cran.r-project.org/). Reporting of results followed the Standards for Reporting Diagnostic accuracy studies (STARD 2015) guideline (13).

### Institutional Review Board (IRB) approval

The IRB Ile-de France III approved the study protocol prior to data collection (approval number 3840-NI) and all subsequent amendments.

## Results

### Participants

Out of 1452 participants enrolled between 19 October and 18 December 2020, one participant did not provide neither nasopharyngeal nor a saliva sample and was excluded from subsequent analyses. Median age of study participants was of 36 years [26-50] and 52% were females (Table 1). Indications for testing and clinical symptoms reported on day of inclusion are detailed in table 1. One to three symptoms were observed in 409/1449 (28%) participants.

**Table 1:** Characteristics of study participants. Results are presented as N(%) or median [interquartile range].

|  | Total<br>N=1451 |
| --- | --- |
| Age, years | 36 [26-50] |
| Females | 755 (52) |
| Medical conditions |  |
| Hypertension | 108 (7) |
| Diabetes mellitus | 31 (2) |
| Hemodialysis | 5 (0.3) |
| HIV infection | 11 (0.8) |
| Solid organ transplant | 19 (1) |
| Contact with a confirmed case | 564 (39) |
| Time from last contact, days | 7 [1-7] |
| Presence of symptoms on day of testing | 571 (39) |
| Time from symptoms onset, days | 3 [2-4] |
| Cough | 292 (20) |
| Headaches | 257 (18) |
| Rhinorrhea | 202 (14) |
| Asthenia | 198 (14) |
| Muscle pain | 177 (12) |
| Fever | 163 (11) |
| Diarrhea | 85 (6) |
| Chills | 69 (5) |
| Anosmia | 62 (4) |
| Shortness of breath | 53 (4) |
| Chest pain | 52 (4) |
| Smoking in the last 24 hours | 293 (20) |
| Consumption of alcohol in the last 24 hours | 320 (22) |
| Consumption of coffee in the last hour | 271 (19) |
| Consumption of coffee in the last 2 hours | 701 (49) |
| Teeth brushing in the last 2 hours | 646 (45) |
| Mouth washing in the last 2 hours | 48 (3) |

### SARS CoV2 positive results

Eight NPS (0.6%) and 24 saliva samples (1.7%) were not appropriately collected and were not analyzed (Appendix Figure 1). In compliant samples, technical failure led to invalid results in 12 (none for nasopharyngeal NAAT and saliva MGI-1, 8 for saliva MGI-2 and 4 for saliva Roche). Overall, 129/1443 (9%) tested positive on nasopharyngeal NAAT: 102/564 (18%) in symptomatic and 27/879 (3%) in asymptomatic participants (Table 2). Detection rates were of 8%, 5%, 13% and 12% for the nasopharyngeal Ag test, saliva MGI-1, saliva MGI-2 and saliva Roche, respectively. As displayed in Figure 1, the overall median Ct value for nasopharyngeal NAAT was of 30.0 [27.6-32.1], with no difference according to the presence of symptoms and timing of testing (p = 0.21) (Figure 1A). The overall median Ct value for saliva NAAT MGI-2 was of 25.1 [22.3-30.3] with significantly lower Ct values in symptomatic participants tested ≤4 days after symptoms onset (23.4 [21.4-26.1]) compared to those tested after 4 days (28.2 [26.1-31.0]) and those with no symptoms (27.2 [24.3-32.6], p<0.001, Figure 1B).

**Table 2:** Number of positive samples (at least one target gene detected) according to the technical procedure: nasopharyngeal Nucleic Acid Amplification Testing (NAAT), nasopharyngeal antigen test and saliva NAAT using three different technical procedures (MGI-1, MGI-2, Roche).

|  |  | Presence of symptoms on day of testing |  |
| --- | --- | --- | --- |
|  |  | Symptoms (n=564) | No symptoms (n=879) |
| Nasopharyngeal NAAT | 129/1443 (9%) | 102/564 (18%) | 27/879 (3%) |
| Nasopharyngeal antigen test | 89/1115 (8%) | 70/464 (15%) | 19/651 (3%) |
| Saliva NAAT / MGI-1 | 17/374 (5%) | 14/174 (8%) | 3/200 (2%) |
| Saliva NAAT / MGI-2 | 167/1334 (13%) | 109/522 (21%) | 58/812 (7%) |
| Saliva NAAT / Roche | 167/1391 (12%) | 116/542 (21%) | 51/849 (6%) |

**Figure 1:**
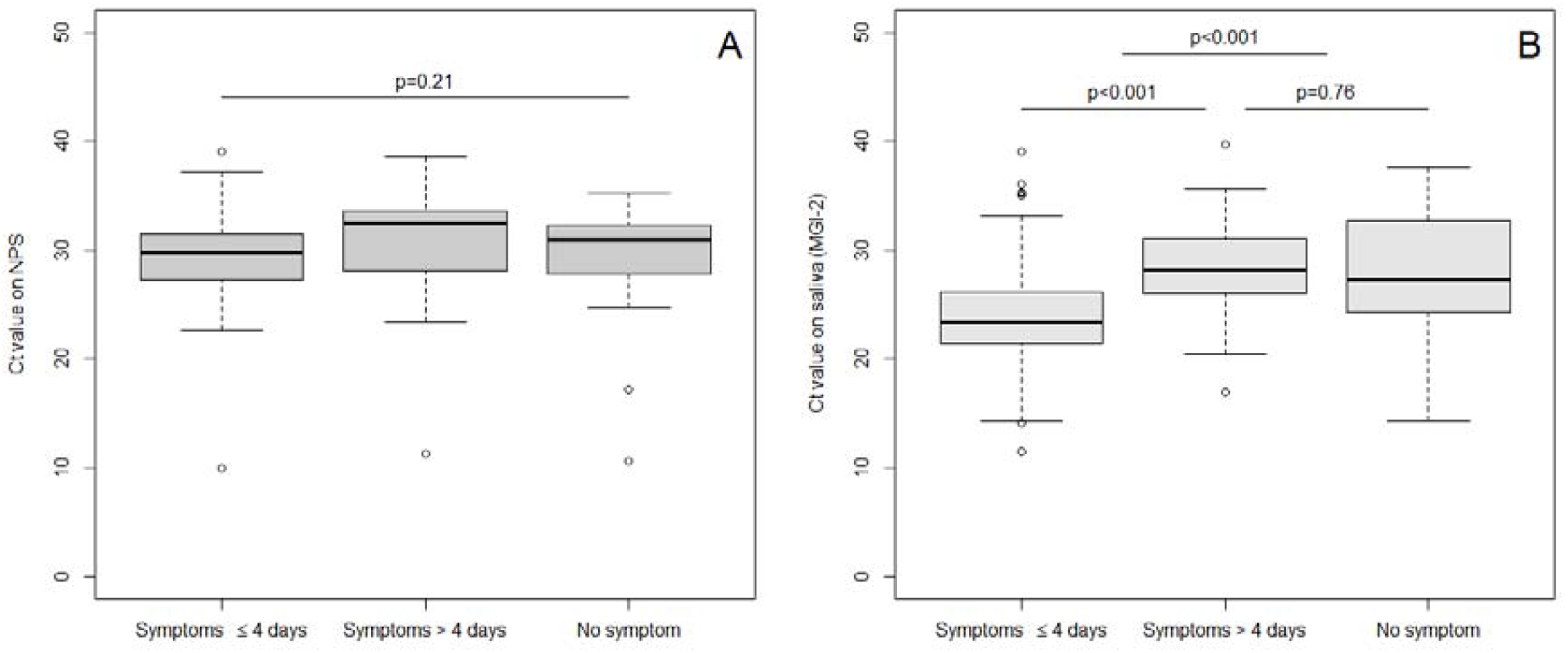
Ct values for nasopharyngeal nucleic acid amplification testing (NAAT) and saliva NAAT (method MGI-2) according to the presence of symptoms and timing of testing after symptoms onset. 1A: nasopharyngeal Ct values, 1B: saliva Ct values (MGI-2)

### Performance of detection of SARS CoV2 infection

Diagnostic accuracy of the nasopharyngeal Ag test and the three NAAT methods on saliva are presented in Table 3. Sensitivity and specificity were similar in symptomatic and asymptomatic participants. Subgroup analyses according to the Ct values are presented in Appendix Table 2. Consumption of alcohol, coffee, food, smoking or teeth brushing within 30 minutes before sampling had no impact on diagnostic accuracy of the three methods (data not shown).

**Table 3:** Diagnostic accuracy of the nasopharyngeal antigen test and saliva NAAT using three different technical procedures (MGI-1, MGI-2, Roche), as compared to the reference standard (nasopharyngeal NAAT, positivity defined as at least one target gene detected), according to the presence of symptoms in study participants.

|  | Total, n | Positive<br>samples, n | Sensitivity<br>(95% CI*) | Specificity<br>(95% CI) |
| --- | --- | --- | --- | --- |
| Nasopharyngeal antigen test | 1109 | 81 | 94% (86-98) | 99% (98-99) |
| Symptoms | 459 | 64 | 95% (87-99) | 98% (96-99) |
| No symptoms | 650 | 17 | 88% (64-99) | 99% (98-100) |
| Saliva NAAT / MGI-1 | 372 | 69 | 23% (14-35) | 100% (98-100) |
| Symptoms | 172 | 60 | 22% (12-34) | 99% (95-100) |
| No symptoms | 200 | 9 | 33% (7-70) | 100% (98-100) |
| Saliva NAAT / MGI-2 | 1328 | 112 | 94% (88-97) | 95% (94-96) |
| Symptoms | 516 | 87 | 95% (89-99) | 94% (92-96) |
| No symptoms | 812 | 25 | 88% (69-97) | 95% (94-97) |
| Saliva NAAT / Roche | 1383 | 120 | 96% (91-99) | 96% (95-97) |
| Symptoms | 535 | 95 | 97% (91-99) | 95% (92-97) |
| No symptoms | 848 | 25 | 92% (74-99) | 97% (95-98) |

\*95% CI: 95% Confidence Interval

We further analyzed correlations between Ct values on nasopharyngeal NAAT and saliva NAAT / MGI-2 (Appendix Figure 2). On NPS and saliva samples, Ct values were moderately correlated (*ρ* = 0.29, p = 0.003), and higher with nasopharyngeal NAAT than saliva NAAT (median differences MIG-2 minus NPS -6.1 [-9.2;-3.7]. On saliva samples, agreement between Ct values with Roche and MGI-2 was moderate (Appendix Figure 3A), Ct values being almost systematically greater with MIG-2 than with Roche (median differences MGI-2 minus Roche -4.2 [-5.2;-2.6], Appendix Figure 3B).

### Analysis of discrepancies

Of 1315 participants who had all three tests (nasopharyngeal NAAT and saliva NAAT with MGI-2 and Roche), 177 had a positive result with at least one technique. The Venn diagram (Appendix Figure 4) displays the number of positive results according to the technique. One hundred and five were positive with all three techniques, 66 were positive only on saliva (including 42 positive with both MGI-2 and Roche), and 5 only on NPS. As displayed on Figure 2, Ct values (MGI-2) were significantly lower in the 105 participants positive on both NPS and saliva (23.7 [21.4-26.1]) than in the 61 positive on saliva only (31.4 [26.1-35.0], p<0.001). Of these 61 participants with positive saliva NAAT not detected with NPS NAAT, 18 (30%) had Ct values ≤28 on saliva.

**Figure 2:**
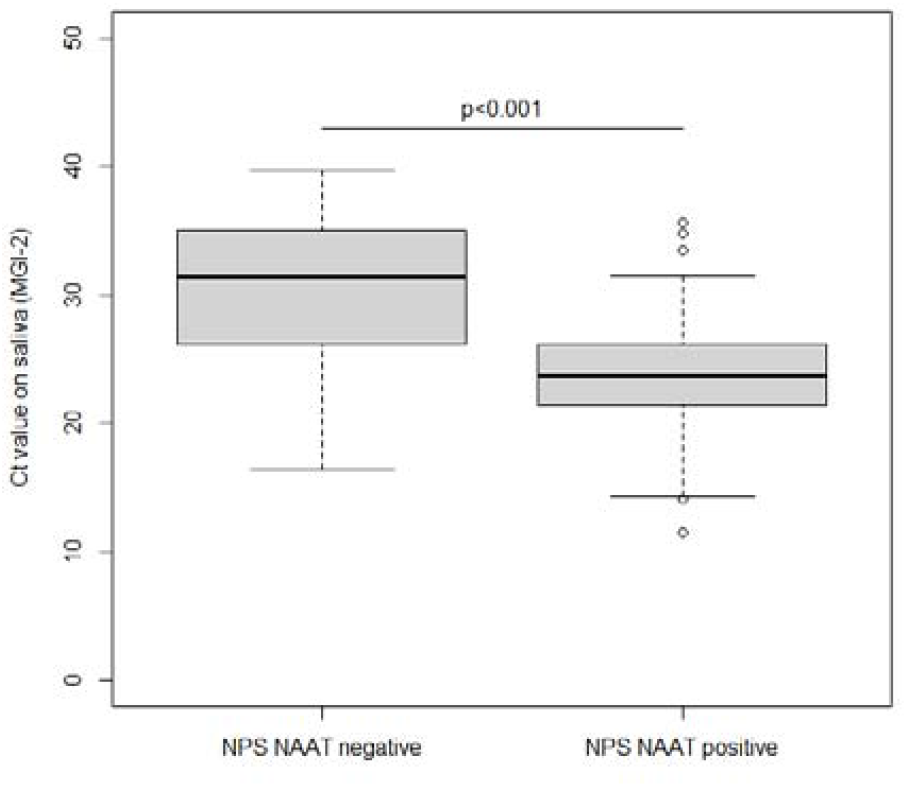
Ct values for saliva NAAT (method MGI-2) according to the result of the nasopharyngeal NAAT.

### Sensitivity analyses

The first sensitivity analysis (reference standard positive if ≥ 2 targets positive on nasopharyngeal NAAT) found similar results (Appendix Table 3) to the main analysis. Results of the second sensitivity analysis (reference standard positive if ≥ 1 target was positive over nasopharyngeal NAAT, saliva MGI-2 or saliva Roche) are presented in Table 4. The overall sensitivity of the nasopharyngeal Ag test decreased to 64% (76% in symptomatic and 40% in asymptomatic participants). The sensitivity of nasopharyngeal NAAT was of 65% (40% in asymptomatic participants) compared to 93% for saliva MGI-2 and 87% for saliva Roche.

**Table 4:** Sensitivity analysis of diagnostic accuracy of the nasopharyngeal antigen test, saliva NAAT with MGI-2 and saliva NAAT with Roche, as compared to the reference standard. The reference standard was considered positive if ≥ 1 target was positive with either the nasopharyngeal NAAT, saliva MGI-2 or saliva Roche.

|  | Total, n | Positive samples, n | Sensitivity (95% CI*) | Specificity† (95% CI) |
| --- | --- | --- | --- | --- |
| Nasopharyngeal antigen test | 1041 | 138 | 64% (55-72) | 100% (100-100) |
| Symptoms | 434 | 91 | 76% (66-84) | 100% (99-100) |
| No symptoms | 607 | 47 | 40% (26-56) | 100% (99-100) |
| Nasopharyngeal NAAT | 1337 | 199 | 65% (58-71) |  |
| Symptoms | 528 | 132 | 77% (69-84) |  |
| No symptoms | 809 | 67 | 40% (28-53) |  |
| Saliva NAAT / MGI-2 | 1317 | 179 | 93% (89-96) |  |
| Symptoms | 511 | 115 | 95% (89-98) |  |
| No symptoms | 806 | 64 | 91% (81-96) |  |
| Saliva NAAT / Roche | 1329 | 191 | 87% (82-92) |  |
| Symptoms | 522 | 126 | 92% (86-96) |  |
| No symptoms | 807 | 65 | 78% (67-88) |  |

\*95% CI : 95% Confidence Interval.
† The specificity for nasopharyngeal NAAT, saliva MGI-2 and saliva Roche is by definition equal to 100% since the three tests are included in the reference standard.

### Acceptability

The median VAS score for pain was of 4 [2-6] for nasopharyngeal sampling and 0 [0-0] for saliva (p<0.001). Median VAS score for simplicity/convenience was of 8 [5-10] for nasopharyngeal sampling and 9 [6-10] for saliva (p<0.001). If sampling had to be repeated in the next days, 127 participants (9%) declared that they would certainly/probably refuse another nasopharyngeal sampling, compared to 43 (3%) for saliva (p<0.001). Main reasons for refusal would be pain (109/127, 86%) for NPS or difficulties to provide the required volume for saliva (6/43, 14%). If participants had the choice between the two sampling methods, 882/1450 (61%) would prefer saliva, 202/1450 (14%) NPS and 366/1450 (25%) would have no preference. The large majority (1423/1451, 98%) estimated to be able to provide a saliva sample self-collected at home.

## Discussion

In this large prospective study in two ambulatory centers, sensitivity of saliva NAAT to detect SARS-CoV-2 varied depending on the pre-processing, amplification and detection procedures, ranging from 23% (with the MGI-1 protocol) to 94-96% (with the MGI-2 and Roche protocols). Specificity was above 95% for all three methods. Diagnostic accuracy of rapid Ag testing was high, with both sensitivity and specificity above 94%. Performances of all tests were similar in symptomatic and asymptomatic participants.

Previous studies on diagnostic accuracy of saliva for detection of SARS-CoV-2 varied greatly in specimen collection, processing protocols, and populations included (9–12). The strengths of our study are the large sample size, including a relatively high number of asymptomatic individuals, and implementation in real-life conditions, on a representative sample of the targeted population, with similar spectrum of disease prevalence and severity. All samples were performed on the exact same day in each participant, and analyzed with the same protocols, for which we provide full details on pre-processing, amplification and detection methods. To avoid any potential dependence to specific devices supply, we used dry tubes to collect saliva samples (14). We also defined criteria for positivity of the different tests and performed sensitivity analyses on these criteria to estimate the impact on diagnostic performances.

Sensitivity of saliva sampling seems similar to that of nasopharyngeal sampling for detection of SARS-CoV-2. Our results further suggest that, with enhanced protocols for pre-processing and detection, saliva sampling could be even more sensitive. The majority of previous studies were lead in symptomatic individuals, and mostly in hospitalized patients. In ambulatory care, sensitivity of saliva NAAT ranges from 70.7% (Confidence Interval: 46.1-96.1%) to 95.7% (93.1-97.5%) (15–23), compared to nasopharyngeal NAAT, and the pooled sensitivity was estimated at 84.5% (73.0%-95.3%) in a recent meta-analysis (10). Most studies did not report details on the presence and dates of clinical signs, the number of genes targeted by NAAT, and only a few were able to obtain paired samples on the first day of presentation, making results difficult to compare. The reason why saliva NAAT was more sensitive than nasopharyngeal NAAT in our study remains unclear. The most straightforward explanation is that our reference standard, nasopharyngeal NAAT, was imperfect, as suggested by others (21). Another hypothesis is that viral RNA is amplified for longer periods from saliva than nasopharyngeal swabs (19,24,25), but the link between detection of viral RNA in saliva and its meaning in terms of infectiousness remains to be clarified.

Our study also highlights that the performances of saliva NAAT strongly depend on processing methods of samples, and illustrates that direct use of NPS methods to saliva could lead to poor performance. In our protocol, the change of lysis buffer in the MGI-2 method increased sensitivity from 23% to 95%. One hypothesis is that the introduction of a significant volume of saliva into lysis buffer, compared to nasopharyngeal secretions collected by swabbing, may alter its properties and decrease efficiency of viral RNA extraction. Our results confirm that SARS-CoV2 RNA detection does not require dedicated reagents for nucleic acids preservation in saliva, and that actually, nucleic acids stabilizing may even affect detection of viral RNA (14). This illustrates that preliminary validation of analytic methods (particularly dilution and lysis buffer) is crucial to provide reliable results on saliva.

Rapid antigen testing showed very high performances in our study, even in asymptomatic individuals. The sensitivity of SARS-CoV2 antigen tests differs significantly according to populations analyzed and antigen tests used. Studies including individuals within 7 days from the onset of symptoms or from exposure to a confirmed case of COVID-19 showed a good concordance with nasopharyngeal NAAT with sensitivity estimates between 77% and 96% (26–28). In symptomatic individuals, antigen tests actually provided a better estimation of infectiousness than nasopharyngeal NAAT (29). Compared to the composite reference (positive results with either nasopharyngeal NAAT or saliva NAAT), the antigen test displayed similar performances than nasopharyngeal NAAT.

This study was limited by the relatively low number of children included. Actually, for many respiratory viruses, diagnostic accuracy of tests vary between adults and children. For SARS-CoV-2, very few studies were specifically designed in this population, but gave encouraging results (30). The ease of collecting non-invasive saliva makes it an attractive specimen for children, particularly for repeated testing. This strategy must however be specifically evaluated before large implementation, especially in young children who may have difficulties to provide the required volume of saliva (31). Second, nasopharyngeal NAAT is an imperfect reference method, which may contribute to overestimate diagnostic performances of alternative tests. Indeed, sensitivity of the nasopharyngeal NAAT was of only 65% (95%CI: 58-71) compared to a composite reference standard (including results of the nasopharyngeal NAAT, saliva MGI-2 and saliva Roche) in the sensitivity analysis. However, NPS is the current reference standard for sampling, and MGI-1 the technique used by the majority of screening centers in France. In this real-life evaluation, we therefore chose to use this method as the reference standard. Similarly, nasopharyngeal sampling was performed by highly trained personnel in our study, which may enhance diagnostic performances of antigen testing.

Saliva sampling is simple, painless, does not require trained personnel, and therefore offers the perspective for self-collection and iterative screening. Its main drawback is to require processing in centralized laboratories, with results available usually within 24 hours. On the other hand, rapid antigen testing provides immediate results, but still needs nasopharyngeal sampling. Considering that diagnostic accuracy seems similar for both methods, their respective advantages should be considered for implementation. Due to high acceptability, saliva NAAT should be considered in community mass screening programs or in situations where iterative screening is necessary. Several simulation studies highlighted the utility of routine iterative noninvasive saliva testing for identification of silent COVID-19 in frontline healthcare workers and prevent outbreaks in healthcare facilities (32,33). By reducing sample-to-answer time, rapid antigen tests are of particular interest to allow immediate identification and prompt isolation of cases. Their point-of-care use makes them particularly useful in primary care practices or in settings where specialized laboratories are not accessible.

### Conclusions

Saliva NAAT and nasopharyngeal Ag testing are reliable alternative strategies to identify SARS-CoV2 in both symptomatic and asymptomatic infected individuals in the ambulatory setting. Their use should be encouraged anywhere it might facilitate screening, tracing and isolation. Dedicated implementation studies are warranted to better assess the optimal respective positioning of these two strategies in the public health response, until rapid, self-collected, reliable tests are available.

## Data Availability

Individual data will not be publicly available, because participants did not provide consent for it.

## Acknowledgments

The authors thank the following members of the scientific committee for insightful comments on the study protocol and results: Bruno Lina, Anne-Geneviève Marcelin, Catherine Paugam-Burtz, Astrid Vabret, Nabil Gastli, and persons involved in data collection: Claire Kappel, Guiseppina d’Anna, Elisabeth Velin, Abdulai Jalloh, Caroline Du Song, Philippine Treluyer, Mathilde Bayle, Lucie Daveau, Marine Hellegouarch, Suzie Zhu, Pénélope Travailleur, Matthieu Kapry, Adeline Huet, Emeline Hermel, Céline Goy, Louise Lavancier.

