## Appendix for "Accuracy of antigen and nucleic acid amplification testing on saliva and naopharyngeal samples for detection of SARS-CoV-2 in ambulatory care"

### **Supplementary appendix**

### I. Methods

#### A. Sampling procedures

As standard protocol, NPS were collected using sterile nylon flocked swab and placed into tubes containing 3 ml of transport medium with guanidium salt and nucleic acid stabilizer (VSM02, Xian Galaxy). The first NPS was sent to the APHP high throughput platform for NAAT as part of routine care (reference method). The second NPS, collected in the second nostril, was used for rapid Ag testing immediately after sampling. The saliva sample was self-collected under supervision of a nurse, after nasopharyngeal swabbing. Participants were asked to salivate in their mouth without clearing their throat and to repeatedly spit into a 50 ml tube up to a maximum of 3 ml. Saliva samples were centralized, frozen in several aliquots at -80°C within 24 hours and stored for analysis.

#### B. Virology methods

An overview of the methods used for pre-processing, extraction and detection are presented in the Appendix Table 1.

##### Nasopharyngeal NAAT

NPS were inactivated at 56°C for 30 minutes. Nucleic acid extraction was performed on 180 µL of NPS with MGIEasy Nucleic Acid Extraction Kit (MGI Tech Co, Shenzhen, China) on a MGISP-960 instrument (MGI Tech Co). SARS Cov-2 RNA amplification was done using TaqPath™ COVID 19 CE IVD RT PCR Kit (Thermo Fisher Scientific, Coutaboeuf, France). This test is a multiplex real-time RT-PCR test intended for the qualitative detection of nucleic acid from SARS‑CoV‑2. The kit contains three primer/probe sets specific to three different SARS-CoV-2 genomic regions (ORF1ab, N and S-genes) and primers/probes for bacteriophage MS2 used as internal control of amplification and extraction. A minimum of one negative control and one positive control was use for each run. Dilution of inactivated SARS-CoV2 cell-culture supernatant (provided by Virology laboratory, Hospices Civils de Lyon) was added once a day in each production line as for external control. The technique provides results expressed as a cycle threshold (Ct) for each gene target. This NAAT procedure is referred as “MGI-1”.

##### Saliva NAAT

Saliva samples were tested with three NAAT procedures: “MGI-1”, “MGI-2” and “Roche”.

In the MGI-1 procedure, after homogenization with a vortex for ﬁve seconds, 150 µl of saliva was mixed with 500 µl of VSM02 buffer, incubated at 56°C for 30 min and then processed for extraction with the procedure used for nasopharyngeal NAAT.

In the MGI-2 procedure, saliva aliquots were thawed, equilibrated to room-temperature and then homogenized with a vortex for ﬁve seconds, and the 300 µl was mixed with 300 µl of NucliSENS® lysis buffer (Biomerieux, Marcy l'Etoile, France) and then extracted with the same procedure used for the NPS. Saliva nucleic acids extracts of MGI-2 were tested with the same RT-PCR procedure than for nasopharyngeal NAAT.

In the Roche procedure, saliva samples were tested using the Roche Cobas® 6800 analyzer and Roche Cobas® SARS-CoV-2 assay (Roche Diagnostics France, Myelan, France) in the virology laboratory in Saint Louis hospital, Paris, France. Thawed saliva samples were equilibrated to room-temperature and vortexed for ﬁve seconds to homogenize the sample and disrupt mucus clots. A 150 µL aliquot of each specimen was transferred a tube containing 500 µL of Cobas omni Lysis Reagent. Samples were vortexed for ﬁve seconds and incubated at room-temperature for 10 min in the Cobas omni Lysis Reagent. The samples were loaded onto the Roche Cobas 6800 analyzer for testing. The Cobas® SARS-CoV-2 test provides fully automated sample preparation (nucleic acid extraction and ampliﬁcation) and RT-PCR-based qualitative detection of SARS-CoV-2 RNA through ampliﬁcation of two genomic target regions, the ORF1ab and E genes. An armored RNA internal control is added to all samples to validate each reaction. A minimum of one negative control and one positive control must be present for each run. Automated data management software assigns test results for all tests.

##### Rapid antigen testing

SARS-CoV-2 antigen testing was performed on one NPS by using Standard Q COVID-19 Ag test (SD Biosensor®, Chuncheongbuk-do, Republic of Korea). Standard Q COVID-19 Ag test is a chromatographic immunoassay for the detection of SARS-CoV-2 nucleocapsid (N) antigen. Directly after sampling, the swab was inserted into the extraction buffer tube. While squeezing the buffer tube, the swab was stirred more than 5 times. The swab was then removed while squeezing the sides of the tube to extract the liquid from the swab. Then 3 drops of extracted specimen were applied to the test cartridge. The result (positive, negative or undetermined if negative control) was read after 15 to 30 minutes according to instructions of the manufacturer.

#### C. Supplementary analysis on fresh saliva

To assess the impact of saliva samples freezing on detection of SARS-CoV2 RNA by RT-PCR, 93 additional consecutive saliva samples were tested twice: the first test was performed on day of sampling and the second test was done 48 hours after storage at minus 20°C. Fresh and frozen samples were tested with the MGI-2 procedure.

Appendix Table 1: Overview of the different NAAT methods used for SARS-CoV-2 detection.

|  | Original sample | Inactivation | Dilution before extraction | Sample for nucleic acid extraction | Technic for nucleic acid extraction | Technic for amplification |
| --- | --- | --- | --- | --- | --- | --- |
| Nasopharyngeal NAAT | Fresh NPS samples | 56°C for 30 minutes |  | 180 µL of NPS | MGIEasy Nucleic Acid Extraction Kit (MGI Tech Co, Shenzhen, China) on a MGISP-960 instrument (MGI Tech Co) | TaqPath™ COVID 19 CE IVD RT PCR Kit (Thermo Fisher Scientific, Coutaboeuf, France) |
| Saliva NAAT / MGI-1 | Fresh saliva samples | 56°C for 30 minutes | 150 µl of saliva + 500 µl of VSM02 buffer | 180 µL of diluted saliva in VSM02 | MGIEasy Nucleic Acid Extraction Kit (MGI Tech Co, Shenzhen, China) on a MGISP-960 instrument (MGI Tech Co) | TaqPath™ COVID 19 CE IVD RT PCR Kit (Thermo Fisher Scientific, Coutaboeuf, France) |
| Saliva NAAT / MGI-2 | Thawed saliva samples | None | 300 µl of saliva + 300 µl of NucliSENS® lysis buffer (Biomerieux, Marcy l'Etoile, France) | 180 µL of diluted saliva in NucliSENS® lysis buffer | MGIEasy Nucleic Acid Extraction Kit (MGI Tech Co, Shenzhen, China) on a MGISP-960 instrument (MGI Tech Co) | TaqPath™ COVID 19 CE IVD RT PCR Kit (Thermo Fisher Scientific, Coutaboeuf, France) |
| Saliva NAAT / Roche | Thawed saliva samples | None | A 150 µL of saliva + 500 µL of cobas omni Lysis Reagent | 400 µL of diluted saliva in cobas omni Lysis Reagent | Roche cobas 6800 analyzer | Roche cobas 6800 analyzer |

### II. Results

#### A. Study participants

Appendix Figure 1: Flow diagram of study participants, according to the Standards for Reporting Diagnostic accuracy studies (STARD 2015) guideline. NPS: Nasopharyngeal Sample, NAAT: Nucleic acid amplification testing; Ag: Antigen

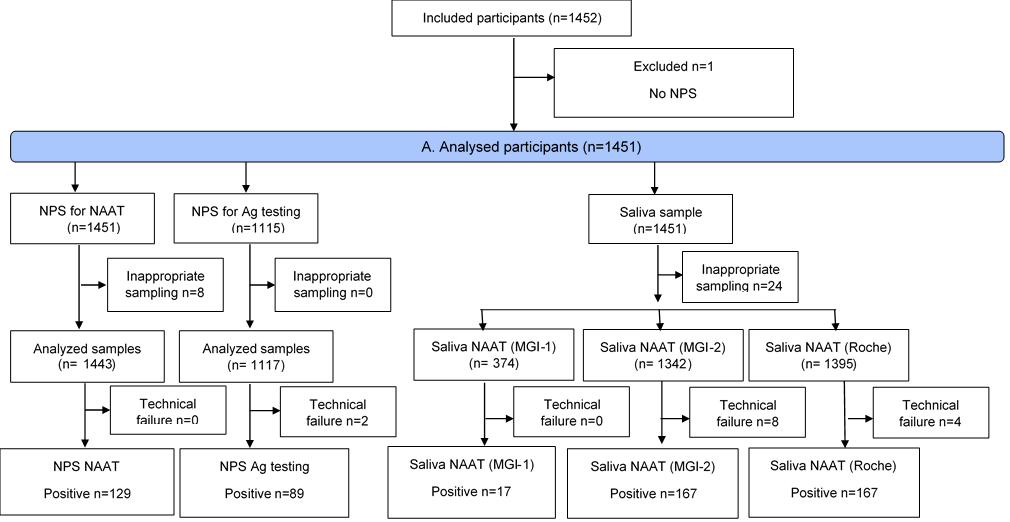

#### B. Subgroup analysis

Appendix Table 2: Subgroup analysis of sensitivity estimates of the nasopharyngeal antigen test, saliva NAAT with MGI-2 and saliva NAAT with Roche, as compared to the reference standard (nasopharyngeal NAAT), according to the Ct values measured with nasopharyngeal NAAT.

|  | Positive samples, n | Sensitivity  (95% CI*) |
| --- | --- | --- |
| Nasopharyngeal antigen test | 81 | 94% (86-98) |
| Nasopharyngeal Ct values ≤ 28 | 28 | 89% (72-98) |
| Nasopharyngeal Ct values > 28 | 53 | 96% (87-100) |
| Saliva NAAT / MGI-2 | 112 | 94% (88-97) |
| Nasopharyngeal Ct values ≤ 28 | 40 | 95% (83-99) |
| Nasopharyngeal Ct values > 28 | 72 | 93% (85-98) |
| Saliva NAAT / Roche | 120 | 96% (91-99) |
| Nasopharyngeal Ct values ≤ 28 | 44 | 95% (85-99) |
| Nasopharyngeal Ct values > 28 | 76 | 96% (89-99) |

*95% CI : 95% Confidence Interval

#### C. Analysis of disprepancies

Appendix Figure 2: Correlation between Ct values on saliva NAAT (Roche) and Ct values on NPS in participants with both tests positive (n=115). 2A: Saliva NAAT performed with the Roche method; 2B: Saliva NAAT performed with the MGI-2 method.

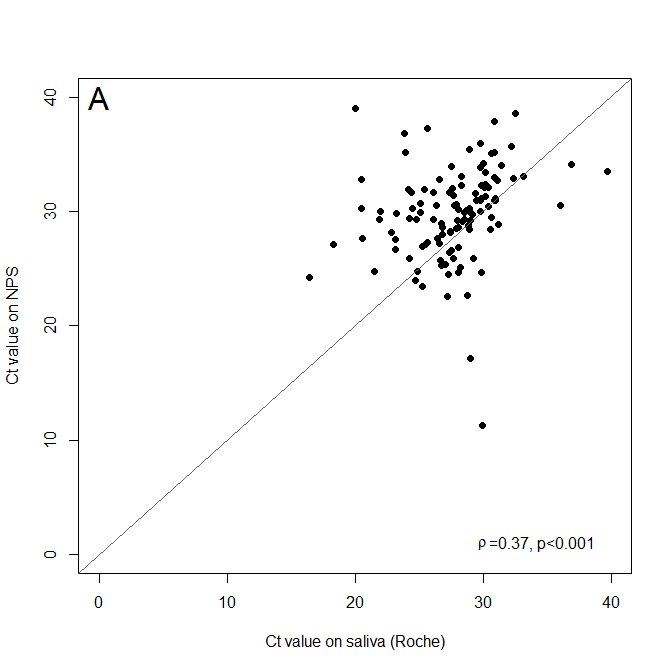

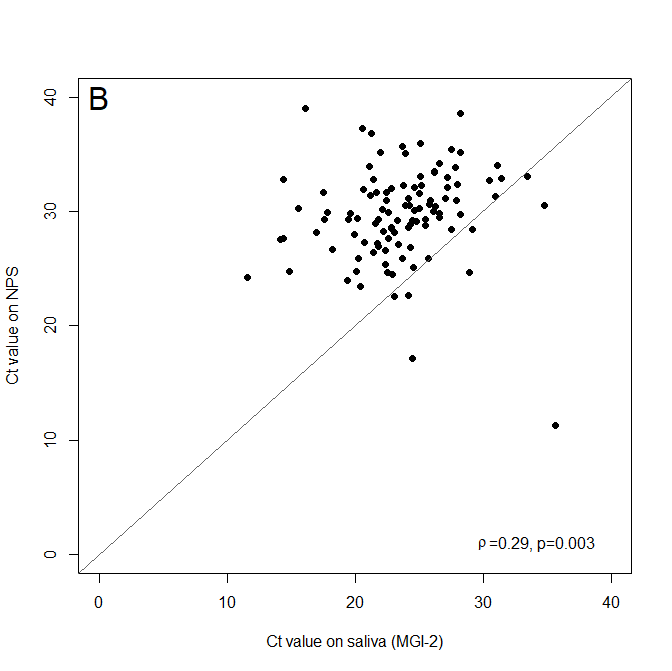

Appendix Figure 3: Agreement between Ct values on saliva with NAAT (MGI-2) and NAAT (Roche) in participants with both tests positive (n=148), 3A: Correlation plot; 3B: Bland-Altman plot. x-axis: average Ct value of the two methods, y-axis: Difference between Ct value with MGI-2 and with Roche method.

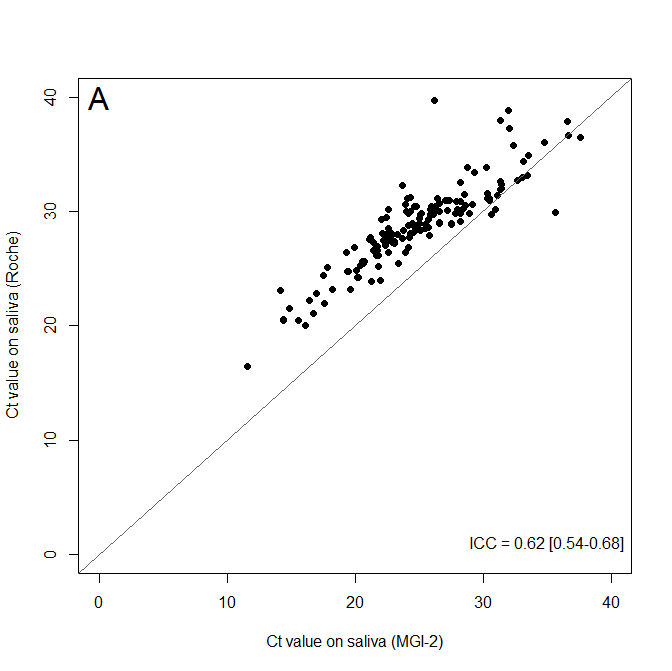

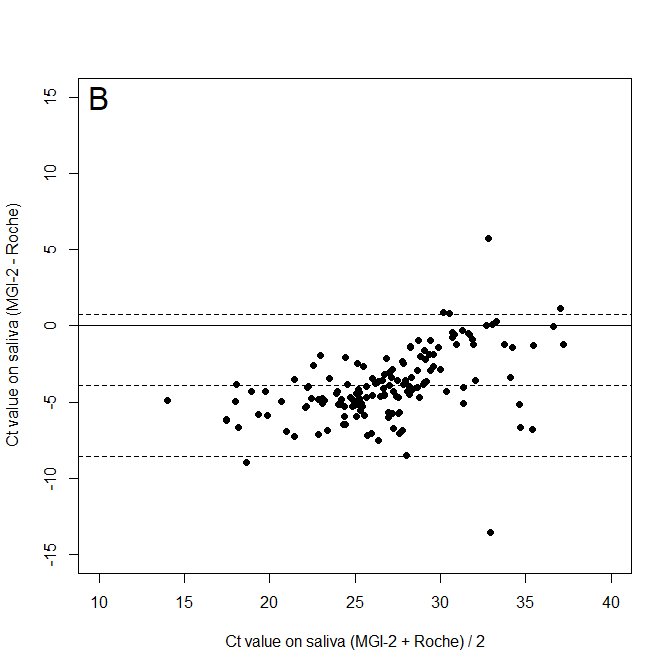

Appendix Figure 4: Venn diagram in 177 participants with at least one test positive (among 1315 having had the 3 tests): NPS NAAT, saliva NAAT (MGI-2) and saliva NAAT (Roche).

NPS: Nasopharyngeal Sample, NAAT: Nucleic acid amplification testing.

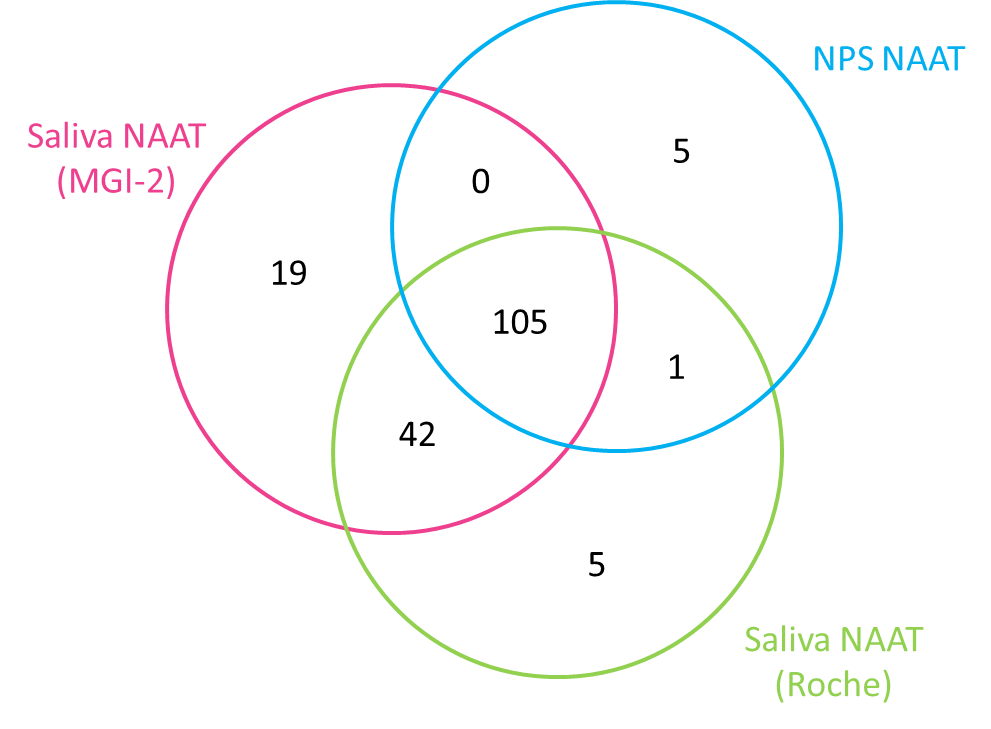

#### D. Sensitivity analyses

Appendix Table 3: Sensitivity analysis of diagnostic accuracy of the nasopharyngeal antigen test, saliva NAAT with MGI-2 and saliva NAAT with Roche, as compared to the reference standard (nasopharyngeal NAAT). The reference standard was considered positive if ≥ 2 targets are positive.

|  | Total, n | Positive samples, n | Sensitivity  (95% CI)* | Specificity  (95% CI) |
| --- | --- | --- | --- | --- |
| Nasopharyngeal antigen test | 1109 | 69 | 97% (90-100) | 98% (97-99) |
| Symptoms | 459 | 57 | 100% (94-100) | 97% (95-98) |
| No symptoms | 650 | 12 | 83% (52-98) | 99% (97-99) |
| Saliva NAAT / MGI-2 | 1328 | 98 | 95% (88-98) | 94% (93-95) |
| Symptoms | 516 | 78 | 97% (91-100) | 93% (90-95) |
| No symptoms | 812 | 20 | 85% (62-97) | 95% (93-96) |
| Saliva NAAT / Roche | 1383 | 106 | 97% (92-99) | 95% (94-96) |
| Symptoms | 535 | 86 | 99% (94-100) | 93% (91-95) |
| No symptoms | 848 | 20 | 90% (68-99) | 96% (94-97) |

*95% CI : 95% Confidence Interval

#### E. Comparison of fresh and frozen saliva samples

Appendix Table 4: Comparison of SARS-CoV2 RNA detection (MGI-2) on fresh and frozen saliva samples. Results was considered positive if ≥ 1 target was detected.

|  |  | Fresh saliva | |  |
| --- | --- | --- | --- | --- |
|  |  | Positive | Negative | Total |
| Fozen  Saliva | Positive | 10 | 1 | 11 |
|  | Negative | 3 | 79 | 82 |
|  | Total | 13 | 80 | 93 |

**Kappa coefficient for overall agreement: 0.81**

Appendix Table 5: Ct values of SARS-CoV2 RNA detection (MGI-2) in samples with positive results on either or both fresh and frozen saliva.

|  | RT-PCR result | |  | Fresh saliva | | | Frozen saliva | | |
| --- | --- | --- | --- | --- | --- | --- | --- | --- | --- |
| Sample number | Fresh saliva | Frozen saliva |  | N gene  Ct value | S gene  Ct value | ORF1ab gene  Ct value | N gene  Ct value | S gene  Ct value | ORF1ab gene  Ct value |
| 001-1178 | + | + |  | 26.15 | 25.65 | 25.41 | 26.92 | 25.9 | 26.78 |
| 001-1196 | + | + |  | 27.19 | 27.43 | 26.87 | 29.07 | 28.75 | 29.32 |
| 001-1198 | + | + |  | 23.29 | 23.34 | 22.89 | 22.44 | 21.48 | 22.24 |
| 001-1211 | + | + |  | 33.51 | 31.66 | 31.19 | 32 | 29.94 | 30.95 |
| 001-1231 | + | + |  | 23.42 | ND* | 23.76 | 23.63 | ND | 25.16 |
| 001-1253 | + | + |  | 21.12 | 19.4 | 18.4 | 21.5 | 19.82 | 20.86 |
| 003-936 | + | + |  | 18.96 | 18.4 | 18.06 | 19.39 | 18.63 | 19.39 |
| 003-947 | + | + |  | ND | ND | 35.77 | ND | ND | 38.84 |
| 003-956 | + | + |  | 23.58 | 22.27 | 22.64 | 23.69 | 22.67 | 23.52 |
| 003-957 | + | + |  | 19.64 | 19.26 | 19.38 | 20.02 | 20.36 | 21.15 |
| 003-943 | - | + |  | ND | ND | ND | ND | 37.97 | ND |
| 001-1207 | + | - |  | 32.59 | 31.21 | 34.84 | ND | ND | ND |
| 001-1216 | + | - |  | 38.2 | 36.38 | 34.74 | ND | ND | ND |
| 001-1250 | + | - |  | ND | 34.55 | 34.55 | ND | ND | ND |

* ND: not detected
